# Risk factors for primary subclavian vein thrombosis in men and women

**DOI:** 10.1101/2025.03.19.25323984

**Authors:** Magdalena Lublin, Jonas Malmstedt

## Abstract

*Introduction:* Primary subclavian vein thrombosis (PSVT) is a rare (2/100.000/year) disorder, affecting predominantly young, healthy individuals. The disease can be idiopathic or effort-induced. The relative importance of risk factors for PSVT is not clarified, sex differences are mostly unknown. Studies show different incidence of idiopathic versus effort-induced PSVT. Aims: To investigate if risk factors differ between men and women with PSVT. The secondary aim was to examine differences regarding time from symptom to diagnosis and treatment between men and women. *Material and* Methods: 135 patients treated for PSVT in Stockholm county 2007–2017 was examined. Data on gender, age, weight, height, smoking, use of oral contraceptives, previous venous thrombotic events (VTE), family VTE, clotting disorders, were collected. Information on whether strenuous muscular activity preceded PSVT debut, and data on time variables where collected. Chi-2-test was used to evaluate differences in categorical data between men and women. Mann-Whitney U-test was used to evaluate time from symptom to first health care contact, diagnosis and treatment. One-way ANOVA was used to adjust for potential confounding variables for time data. Results: More men than women had effort-induced PSVT debut (p=0.006). Men had longer patient delay than women (p=0.064). The difference remained significant after adjustment for strenuous muscular activity. Conclusions: It was more common with effort-induced PSVT amongst men than women. Patient-delay was longer for men than women. Because of the study design, there was no control group, more research is needed to elucidate if causation is present.

## Introduction

Upper-extremity deep vein thrombosis (UEDVT) is a rare condition that accounts for about 10% of all deep vein thrombosis (DVT) (1). UEDVT consists of a broad spectrum of thrombosis in several different segments of veins in the upper extremity. The most common location for thrombosis within UEDVT is the subclavian vein followed by the axillary vein and the brachial vein (2).

The risk factors for UEDVT differ from the risk factors for DVT. Compared to patients with DVT, patients with UEDVT are more likely to be younger, leaner and lack history of DVT (1,3). UEDVT occurs spontaneously or is associated with pacemaker use, central vein catheter (CVC) use or malignancy (4). Accordingly, UEDVT can be divided into primary and secondary forms as presented in Figure 1. Primary subclavian vein thrombosis (PSVT) lacks known external or underlying causes, whereas secondary UEDVT is always associated with an external or underlying cause (4). PSVT is a rare disease, affecting predominately young individuals in their 20-ies and 30-ies, with potentially life-long complications if untreated, which makes it an important disease for clinicians to recognize and treat appropriately (4,5). Secondary UEDTV is more common than PSVT (4). Though secondary UEDTV has a diverse etiology, it is always associated with external factors e.g. malignancy, CVC, pacemaker cables, clotting disorders, malignancy or trauma (4). The most common underlying cause of secondary UEDVT is the use of CVC. A majority of patients with secondary UEDTV has mild, or no related symptoms or signs, which is not the case for PSVT (1,6).

**Figure.1.**
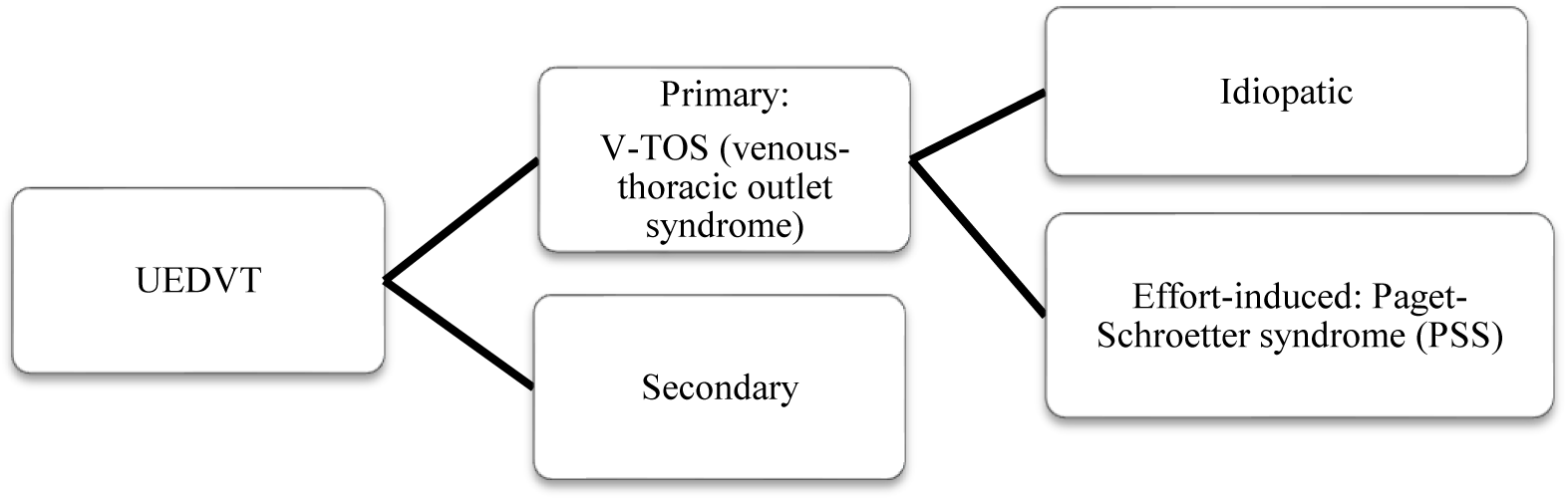
Basic classification of upper extremity deep vein thrombosis entities (UEDVT)

### Pathogenesis for PSVT

The subclavian vein is vulnerable to injury by compression as it passes between the clavicle and the first rib of the thoracic outlet (5). In most cases the etiology of the injury is repeated micro-trauma to the subclavian vein due to an anatomically narrow outlet, thoracic outlet syndrome (TOS) resulting in damage to the subclavian vein endothelium and ultimately subsequent thrombosis (7–9). Due to abnormal anatomy or hypertrophy of the muscles in this area (m. subclavius and m. scalenus anterior), surrounding structures compress the subclavian vein and the condition is also known as venous thoracic outlet syndrome (V-TOS) (5,9). V-TOS can either have idiopathic or effort-related causes (Figure 1). Idiopathic cases lack known cause or trigger factor whilst the effort-related cases, also called Paget-Schroetter syndrome (PSS) is related to strenuous arm work (10). The condition is worsened by an elevated arm position during exercise or strenuous work since the vein is pinched between the clavicle and the first rib. The micro-trauma damages the vessel and a local inflammation including endothelial dysfunction is induced, which in turn can generate fibrosis with thickening of the intima of the vein. This leads to a narrower lumen of the vessel with reduced blood flow and over time occlusion may occur (5,11). Approximately 50% of the PSVT cases are effort-induced (PSS), typically generated by strenuous movements of the arm and shoulder involving abduction and externally rotation, e.g. swimming, painting or weight lifting (11). Since PSVT predominately affects young individuals without traditional risk factors for venous thrombosis, it differs from an UEDVT diagnosis, and there is a need for further understanding of the disease (4).

### Clinical presentation

The clinical presentation of PSVT usually includes sudden onset of swelling, heaviness, pain and discoloration of the affected arm, sometimes accompanied by superficial dilated veins on the chest, shoulder or neck (5). The mean age presentation for PSVT is the early thirties according to a review by Illig et al. and among these patients 60-80% had reported vigorous exercise or activity involving the upper extremities (5). This should be compared to the study by Aziz from 1986 where the incidence of PSS among PSVT patients was approximately 50% (11). To be noted is that Isma et al. reported an effort-induced etiology for PSVT in only 2% of their 63 examined cases (12). Accordingly, the significance of strenuous work prior to debut of PSVT is not entirely known.

### Diagnostics and treatment strategies

Duplex ultrasound (DUS) is the initial diagnostic tool for PSVT. It is a tool with high sensitivity 78-100% and high specificity 82-100% (13). When the result of DUS is negative or uncertain, contrast venography or computed tomography (CT) is used.

The aim of PSVT treatment is relief of symptoms, halt thrombus propagation, prevention of recurrent thrombosis and prevention of complications i.e. pulmonary embolism (PE) and post thrombotic syndrome (PTS). Since randomized controlled trials on treatment strategies are lacking, guidelines are based on data from retrospective studies and clinical experience, and there is need for randomized controlled trials (5). There are three principal treatment strategies for PSVT: conventional treatment e.g. anticoagulation alone, active treatment e.g. early catheter-directed thrombolysis (CDT) combined with early decompressive surgery and lastly conventional treatment with delayed surgery. The foundation of conventional PSVT treatment is anticoagulation. The primary aim is to prevent pulmonary embolism (PE) and thrombus expansion. Anticoagulation alone, e.g. use a vitamin K antagonist (VKA) for more than 3 months after debut of thrombosis is suggested by the guidelines of American College of Chest Physicians (CHEST) and from the American Heart Association (14). Though it has been proposed by several studies, anticoagulation alone is insufficient to prevent recurrent thrombosis and remaining symptoms (15–17). Active treatment consists of early CDT combined with anticoagulation. To achieve best results, thrombolysis should be performed no later than 14 days from onset of symptoms (18) otherwise the chance of a complete thrombolysis is poor (15). Alongside with CDT most centers provide decompressive surgery to treat the underlying cause of PSVT (16,17,19,20). The surgical procedure involves removal of the anterior part of the first rib, and removal of fibrotic tissue surrounding the affected vein. This treatment regime is not suggested by CHEST and American Heart Association, but it has showed promising result by several authors (17,19,20). For patients, unable to get interventional treatment within 14 days, there is the third option. Anticoagulation combined with late surgery. As stated above there are conflicting opinions on which treatment strategy to use in PSVT treatment for optimal benefit.

### Complications

A well-known potentially fatal complication to DVT is PE. However, patients with PSVT have a lower risk to develop PE than patients with DVT in lower extremities or patients with secondary UEDVT (21). Kooji et al. showed that the risk for PE among patients with secondary UEDVT was doubled compared to patients with PSVT (21).

PSVT has minimal long-term sequelae, if treated correctly, but is associated with significant long-term morbidity if ignored (5). Post-thrombotic syndrome (PTS) is the most common chronic complication to venous thrombosis. PTS is characterized by chronic manifestations of venous insufficiency such as swelling, edema, pain and heaviness in the affected limb (22). The pathogenesis of PTS is venous hypertension caused by outflow obstruction in the damaged vein. There are no certain estimates of incidence of PTS in PSVT patients, but it has been reported among 7-46% of patients with preceding UEDVT (22). PTS seems to be less common among secondary UEDVT patients, and more common among patients with PSVT (23). Though the true incidence of PTS remains uncertain due to lack of proper data and lack of diagnostic standard.

### Epidemiology and predisposing factors

Since UEDVT is rare and PSVT is even more rare, there is only one small population-based study made on UEDVT, giving an incidence of approximately 3.6/100 000 individuals per year. The study included both primary and secondary subclavian vein thrombosis i.e. incidence for UEDVT. According to the same study the yearly incidence of PSVT was approximately 2/100,000 inhabitants (3). The information on predisposing factors for PSVT is limited. Though Sir Paget in 1875 and Von Schroetter in 1884 reported effort-induced subclavian vein thrombosis, and Rudolph Matas described an association between upper extremity venous thrombosis and strenuous work in the year 1934, there is still remaining uncertainty about the connection between PSVT and physical strenuous work (10,24). The reported incidence of PSS among PSVT patients ranges from 2 up to 80 percent (3,5,11,25). Isma et al. concluded that thrombophilia (such as Factor V-, prothrombin- and Leiden-mutation) constitutes a predisposing factor for UEDVT, whereas the role of oral contraceptives remains unclear according to the same study (12). Additionally, the sex-specific incidence of PSVT is not entirely known since studies have suggested both male and female dominance (17,20,25). Neither is it known whether patient characteristics differ between men and women. Consequently, there is uncertainty about which factors affect the risk of getting PSVT. One study that investigated the role of thrombophilia and oral contraceptives in patients with PSVT concludes that oral contraceptives cannot be considered a risk factor unless combined with thrombophilia. It also concluded that the risk of getting PSVT was higher among individuals with inherited thrombophilia than in those without (25). Though a positive history of family VTE (venous thrombus embolism) was not significant among PSVT patients compared to the control group (25). As stated above PSVT unlike DVT predominately affects young individuals who are lacking traditional risk factors for venous thrombosis, such as high age, previous VTE, obesity and comorbidities i.e. heart failure and/or vascular disease (4). There are no studies investigating potential differences between men and women according to time from symptom to treatment, i.e. patient delay and doctors delay. Taken together there is a gap of knowledge regarding both incidence, sex differences and the distribution of risk factors for PSVT.

### Aims

The first aim was to investigate if potential risk factors differed between men and women with PSVT. The secondary aim was to examine whether there were any differences regarding time from symptom to diagnosis and treatment between men and women.

## Materials and Methods

This was a retrospective cohort study. The study population was all known PSVT patients treated with CDT, CDT and early surgery, anticoagulation and late surgery, or anticoagulation alone in Stockholm County. The study design was selected since this is a rare disease, and the study design therefore included all known patients with PSVT in Stockholm County between 2007 and 2017. This is part of Magdalena Lublins Degree project 30 credits, spring 2018 at the Study Program in Medicine at Karolinska Institutet.

### Study population

The study population consisted of 135 patients, treated for PSVT in Stockholm county during January 2007 to February 2017. The inclusion criteria were DUS and/or CT-verified diagnosis of thrombosis in the subclavian vein, i.e. all PSVT patients. Patients with secondary UEDVT were excluded. Secondary UEDVT was defined as UEDVT associated with: malignancy, recent trauma (e.g. clavicle fracture), previously known clotting disorder with earlier vein thrombosis at other location, CVC-use, pacemaker-use, recent critical illness or pregnancy.

**Figure 2.**
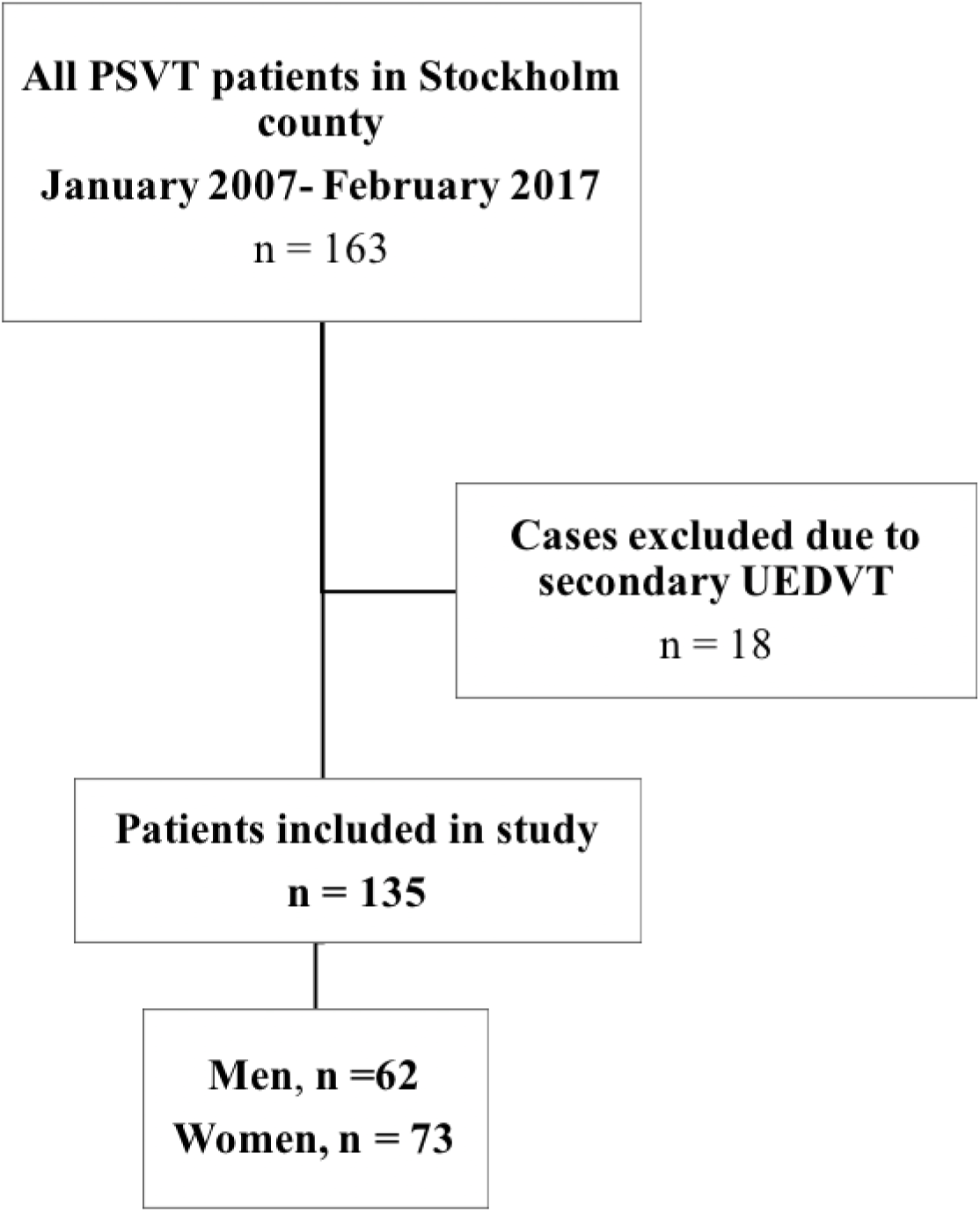
Flowchart displaying patient selection. Secondary UEDVT was defined as UEDVT associated with malignancy, recent trauma (e.g. clavicle fracture), previously known clotting disorder with earlier vein thrombosis at other location, CVC-use, pacemaker-use, recent critical illness or pregnancy. Abbreviations: PSVT= primary subclavian vein thrombosis, UEDVT= upper extremity deep vein thrombosis, N = number of subjects, CVC=central venous catheter.

### Data collection

Patients treated for PSVT in Stockholm county 2007 to 2017 were identified from a prospective registry database at the two units for vascular surgery in Stockholm (Karolinska Universitetssjukhuset and Södersjukhuset). The data were compiled into a raw database, and entered into EpiData® software. Some of the dataset had already been compiled by a previous student, while some of the data had been collected by us. All data were collected through medical records. The collected data included patient demographics such as age, gender, height, weight, occupation, handedness and affected arm. Data on potential predisposing factors included preceding strenuous activity, smoking, use of oral contraceptives (OC), family history of VTE, previous VTE, known clotting disorders and info on regular work/sports with potential to affect the arm. Strenuous activities were defined as muscular activities involving the affected arm, preceding debut of PSVT, of repetitive or strenuous character; including heavy lifting, prolonged above-shoulders extension or abduction of the arm. Smoking was defined as ongoing smoking at time of PSVT diagnosis. OC was defined as ongoing use of oral contraceptives at time of PSVT diagnosis. A positive history of family VTE was considered when objectively documented episodes among at least one first- or second degree relative where reported. Previous VTE was defined as objectively documented episode of VTE prior to debut of PSVT. Known clotting disorders were defined as positive factor II- or factor V-mutation, positive anti-thrombin, protein-C, protein-S deficiencies or antiphospholipid antibodies (lupus anti-coagulant, anti-cardiolipin, or anti-Beta-2GP1), detected after PSVT-diagnosis. The variable work/sports/activity affecting arm in everyday life were defined as any work with extended arm position e.g. professional driver, painter, builder, and any regular sports/activity, -i.e. exercised once a week or more, including extensive arm use e.g. swimming, dancing, playing hockey, playing tennis etc.

Additionally, data on time delays were collected. Patient delay was defined as time from first symptom to first health care contact. Doctors delay was defined as time from first health care contact to time for diagnosis. Total delay was defined as time from debut of symptom to time for diagnosis was defined. Intervention-CDT-delay was defined as time from debut of symptom to start of CDT. Intervention-surgery-delay was defined as time from debut of symptom to start of surgery. Alongside, data on treatment details were collected including data on treatment strategies. The patients were grouped in two different treatment groups based on time to intervention: acute group and conservative group (the rest of the patients). The acute group got CDT and surgery within 14 days from onset of symptom debut. The conservative group got conservative treatment i.e. no CDT or surgery.

### Outcome

The primary outcome was the difference in proportions of certain risk factors for PSVT in men compared to women. The secondary outcome was sex differences in delay from first symptom to healthcare contact, diagnosis and treatment

### Statistical analysis

Descriptive statistics for baseline characteristics are presented separately for men and women. The normal distribution was tested by inspection of normal Q-Q plots and by Shapiro-Wilks test. Continuous variables with non-normal distributed data were described with median and inter-quartile range (IQR), while normal-distributed data were described by mean and standard deviation (SD). A Mann-Whitney-U test was used to test for sex differences in median age at PSVT debut. For continuous data with normal distribution T-test was used. We also categorized debut age of PSVT into three groups based on tertiles in order to compare the distribution of age in men and women. We used two-sided Chi-2-square-tests to evaluate the frequency of categorical variables in women compared to men. A p-value <0.05 was regarded as significant. The frequency of each possible risk- or predisposing factor in men compared to women was first evaluated in univariate analysis with a Chi^2^-test. We used ANOVA to account for potential confounding in time to healthcare contact, diagnosis and treatment between men and women. Data were collected via EpiData® software, version 4.0.2.49 (Comprehensive Data Management and Basic Statistical Analysis System. Odense. Denmark) and analyzed in SPSS version 22 software (IBM, USA).

### Ethical considerations

This study required accessing and reading of medical records and an ethical permit was considered necessary. The study consisted of data on patients who had already been clinically treated therefore there was no danger for physical harm or danger regarding the enrolled patients. The ethical issue was instead accessing and reading of patients’ medical records, since patients were not asked for their permission to this access which might be seen as an intrusion of the personal integrity. The study itself did not include any personal data such as personal numbers or names, all data were presented on a statistically group level, which makes it impossible to track to any participating individual. All PSVT patients in Stockholm county i.e. all participants in this study had been registered in Swedvasc (quality index of Swedish vascular surgery). Patients were asked to fill in a quality of life form, not included in this study, but part of a bigger project of which this study was a part. Upon the registration, patients were informed about the study of which this was part and it was made clear that participation was voluntary and that data would be treated confidentially by authorized personnel. The submission of the form was considered agreement to participate in the study.

As stated above, the disadvantage of this study from an ethical point of view is the intrusion of patient’s personal integrity. The advantages of this study would be to get valuable information of the unusual diagnosis of PSVT and its predisposing factors. Our total evaluation is that the advantages of this particular study out-rules the possible intrusion of privacy for the participating patients. The benefit of getting knowledge of possible risk factors and potential gender differences is considered greater than the inconvenience of personal integrity compromises that the study may apply. An ethical permit was approved by the Regional Ethic Review Board in Stockholm 2017-01-18 (DNR 2016/2484-31/2).

## Results

The baseline characteristics for the study population are presented in Table 1. There were slightly more women (54%) than men among the PSVT patients over all. Among the men there was a majority who had been engaged in strenuous muscular activity preceding debut of PSVT, this difference was significant compared to the women included. Generally, the right arm was more commonly affected than the left in both men and women. The mean BMI amongst all patients was 23.7. A majority of the men compared to women were engaged in regular work, sports, or activity affecting the arm, though the difference was not significant.

**Table 1.**
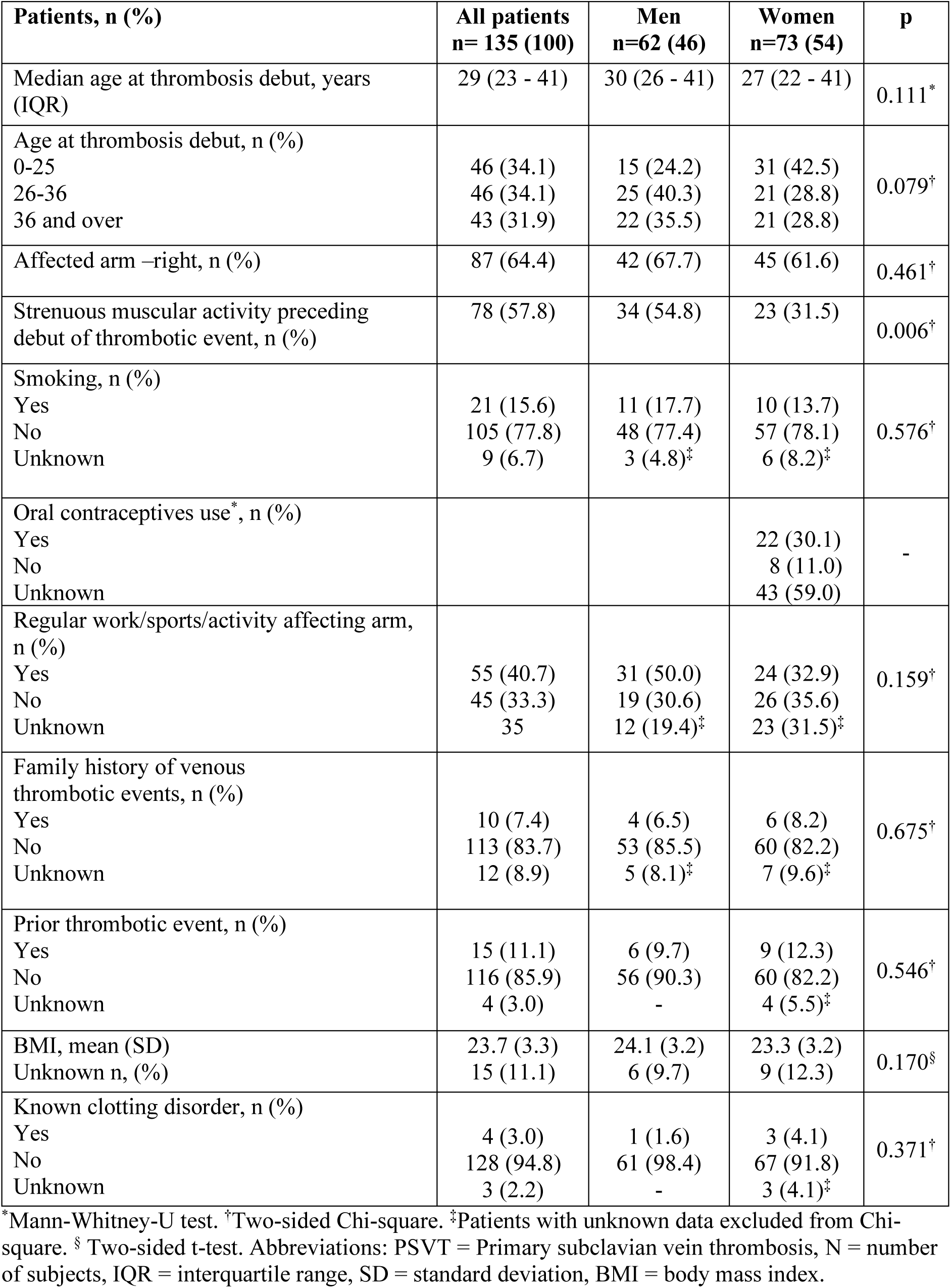
Baseline characteristics for patients with PSVT divided into men and women. Characteristics of the study population for all PSVT patients in Stockholm county year 2007-2017.

| **Patients, n (%)** | **All patients**<br>**n= 135 (100)** | **Men**<br>**n=62 (46)** | **Women**<br>**n=73 (54)** | **p** |
| --- | --- | --- | --- | --- |
| Median age at thrombosis debut, years (IQR) | 29 (23 - 41) | 30 (26 - 41) | 27 (22 - 41) | 0.111* |
| Age at thrombosis debut, n (%) |  |  |  | 0.079† |
| 0-25 | 46 (34.1) | 15 (24.2) | 31 (42.5) |  |
| 26-36 | 46 (34.1) | 25 (40.3) | 21 (28.8) |  |
| 36 and over | 43 (31.9) | 22 (35.5) | 21 (28.8) |  |
| Affected arm –right, n (%) | 87 (64.4) | 42 (67.7) | 45 (61.6) | 0.461† |
| Strenuous muscular activity preceding debut of thrombotic event, n (%) | 78 (57.8) | 34 (54.8) | 23 (31.5) | 0.006† |
| Smoking, n (%) |  |  |  | 0.576† |
| Yes | 21 (15.6) | 11 (17.7) | 10 (13.7) |  |
| No | 105 (77.8) | 48 (77.4) | 57 (78.1) |  |
| Unknown | 9 (6.7) | 3 (4.8)‡ | 6 (8.2)‡ |  |
| Oral contraceptives use*, n (%) |  |  |  | - |
| Yes |  |  | 22 (30.1) |  |
| No |  |  | 8 (11.0) |  |
| Unknown |  |  | 43 (59.0) |  |
| Regular work/sports/activity affecting arm, n (%) |  |  |  | 0.159† |
| Yes | 55 (40.7) | 31 (50.0) | 24 (32.9) |  |
| No | 45 (33.3) | 19 (30.6) | 26 (35.6) |  |
| Unknown | 35 | 12 (19.4)‡ | 23 (31.5)‡ |  |
| Family history of venous thrombotic events, n (%) |  |  |  | 0.675† |
| Yes | 10 (7.4) | 4 (6.5) | 6 (8.2) |  |
| No | 113 (83.7) | 53 (85.5) | 60 (82.2) |  |
| Unknown | 12 (8.9) | 5 (8.1)‡ | 7 (9.6)‡ |  |
| Prior thrombotic event, n (%) |  |  |  | 0.546† |
| Yes | 15 (11.1) | 6 (9.7) | 9 (12.3) |  |
| No | 116 (85.9) | 56 (90.3) | 60 (82.2) |  |
| Unknown | 4 (3.0) | - | 4 (5.5)‡ |  |
| BMI, mean (SD) | 23.7 (3.3) | 24.1 (3.2) | 23.3 (3.2) | 0.170§ |
| Unknown n, (%) | 15 (11.1) | 6 (9.7) | 9 (12.3) |  |
| Known clotting disorder, n (%) |  |  |  | 0.371† |
| Yes | 4 (3.0) | 1 (1.6) | 3 (4.1) |  |
| No | 128 (94.8) | 61 (98.4) | 67 (91.8) |  |
| Unknown | 3 (2.2) | - | 3 (4.1)‡ |  |

\*Mann-Whitney-U test. †Two-sided Chi-square. ‡Patients with unknown data excluded from Chi-square. § Two-sided t-test. Abbreviations: PSVT = Primary subclavian vein thrombosis, N = number of subjects, IQR = interquartile range, SD = standard deviation, BMI = body mass index.

The median age for symptom debut of PSVT for all included patients in the study was 29 years (range 13-64 years) (Table 1.). The population was divided in three age groups according to symptom debut based on tertiary range. There was a predominance of women in the youngest group (0-25 years) compared to men. The women in this group represented 42.5% compared to 24.2% of men (Chi2=5.074, df=2, p=0.079). When comparing men and women aged 0-25 years with those older than 25 years, the dominance of women in the youngest age group was apparent (Chi2=4.983, df=1, p=0.026). There were no other significant differences in the baseline characteristics in men compared to women.

Among all PSVT patients, the total delay (time from symptom debut to diagnosis) was longer for men, compared to women. The patient delay (time from symptom debut to first health care contact) was also longer for men than women (Table 2).

**Table 2.** Time from symptom debut to first health care contact and diagnosis for all PSVT (primary subclavian vein thrombosis) patients, divided by men and women.

|  | **All patients**<br>**(n=135)** | **Men**<br>**(n=62)** | **Women**<br>**(n=73)** | **p\*** |
| --- | --- | --- | --- | --- |
| Total delay (time from first symptom to diagnosis), median, days (IQR) | 4 (2-8) | 5.5 (2-10.8) | 3 (1-7) | 0.054 |
| Patient delay (time from first symptom to first health care contact, median, days (IQR) | 3 (1-7) | 3.5 (1-7.8)<br>(2 missing) | 1.5 (0.3-6)<br>(5 missing) | 0.068 |
| Doctors' delay (time from first health care contact to diagnosis), median, days (IQR) | 0.0 (0.0-2.8) | 1.0 (0.0-2)<br>(2 missing) | 1 (0.0-3.8)<br>(5 missing) | 0.408 |

\*Mann-Whitney U-test. Abbreviations: PSVT = Primary subclavian vein thrombosis, N = number of subjects, IQR = interquartile range.

In the acute group (patients treated with CDT and VKA within 14 days from symptom debut), patient delay was longer for men compared to women. There was no significant difference for total delay in the acute group (Table 3). There were no significant sex differences in doctors delay or intervention delay, neither for lysis (CDT) or surgery.

**Table 3.** Time from symptom debut to first health care contact, diagnosis and intervention for all *acute* PSVT patients, divided by men and women.

|  | **All acute patients**<br>**(n=74)** | **Men**<br>**(n=32)** | **Women**<br>**(n=42)** | **p\*** |
| --- | --- | --- | --- | --- |
| Total delay (time from first symptom to diagnosis), median, days (IQR) | 3 (1-7) | 4 (1.3-7.8) | 3 (1-5) | 0.269 |
| Patient delay (time from first symptom to first health care contact), median, days (IQR) | 1 (0.0-4.3) | 3 (1-6.5) | 1 (0.0-3.0) | 0.064 |
| Doctors' delay (time from first health care contact to diagnosis), median, days (IQR) | 0.00 (0.0-1.0) | 0.0 (0.0-1.0) | 0.0 (0.0-1.3) | 0.546 |
| Intervention delay: lysis (CDT), median, days (IQR) | 5 (2-9) | 5 (2-9)<br>(3 missing) | 5.5 (2.8-8.3)<br>(4 missing) | 0.804 |
| Intervention delay: surgery, median, days (IQR) | 14 (9-22) | 13 (8.3-21.8)<br>(4 missing) | 15 (9.3-26)<br>(10 missing) | 0.309 |

\*Mann-Whitney U-test. Acute group = Treatment with CDT and VKA within 14 days from symptom debut. Abbreviations: PSVT = Primary subclavian vein thrombosis, N = number of subjects, IQR = interquartile range, CDT = catheter-directed thrombolysis, VKA = vitamin K antagonist.

Factors significantly associated with sex e.g. patient delay and muscular strenuous activity was further analyzed in a multivariate ANOVA model. Due to extreme outlying values, four patients with >70 days patient delay, were excluded from the analysis. Results are presented in Figure 3. The difference in patient delay between male and female remained significant after adjustment for strenuous muscular activity p=0.039 (ANOVA, F=4.4, df=3). Both men and women had shorter patient delay with strenuous muscular activity preceding PSVT debut.

**Figure. 3.**
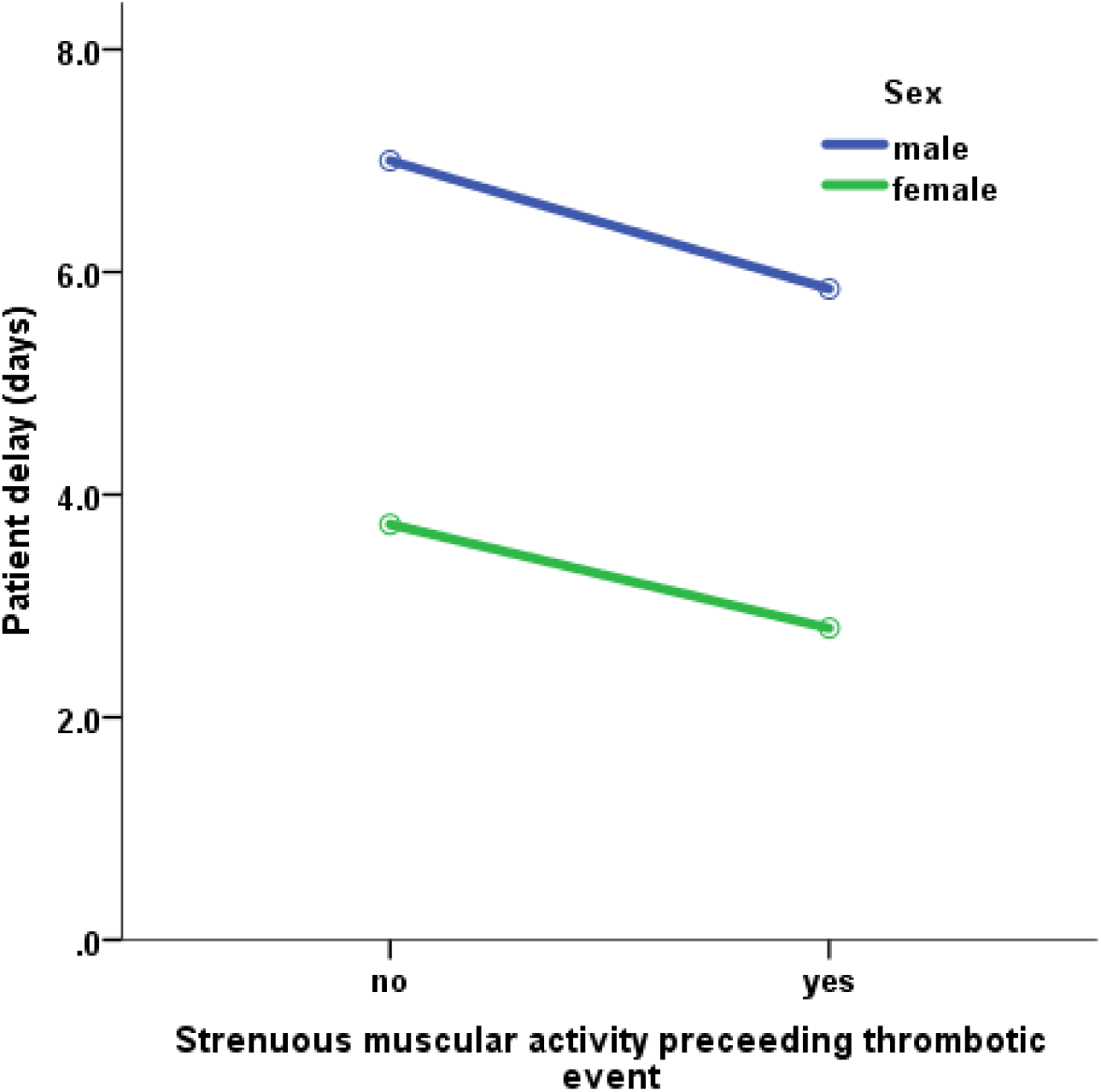
Patient delay for men compared to women with or without strenuous muscular activity preceding PSVT debut. Patient delay defined as time from first symptom to first health care contact presented as median days. PSVT = primary subclavian vein thrombosis.

Doctors delay was analyzed using the same method. Results are presented in Figure 4. There was a significant interaction between sex and strenuous muscular activity (p= 0.040) (ANOVA F=4.3, df=1). Men with preceding strenuous muscular activity had longer doctors delay than men without. Women with preceding strenuous muscular activity had instead shorter doctors delay than women without.

**Figure. 4.**
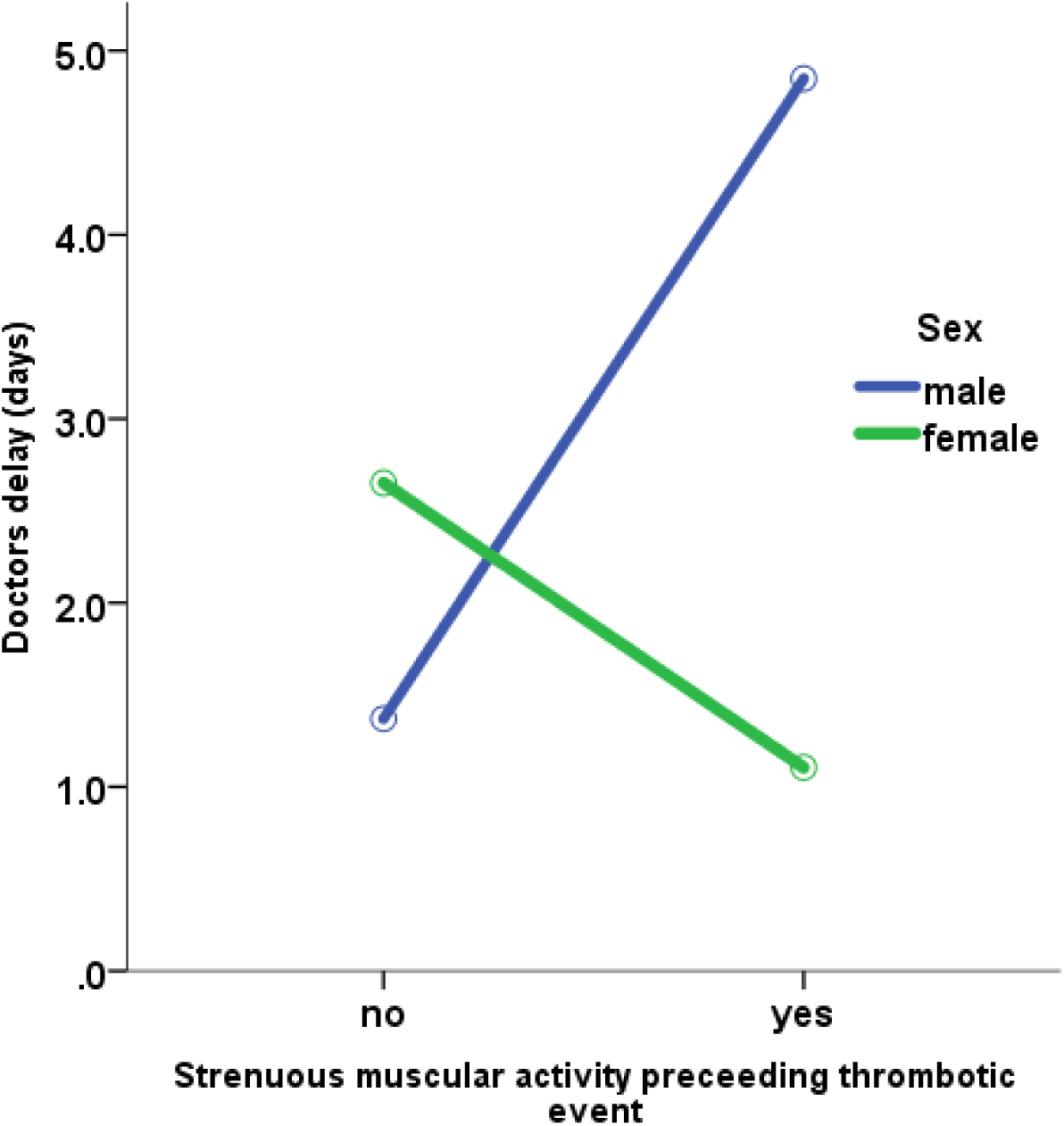
Doctors delay for men compared to women, with or without strenuous muscular activity preceding PSVT debut. Doctors delay defined as time from first health care contact to diagnosis presented as median days. PSVT = primary subclavian vein thrombosis.

## Discussion

To our knowledge this is the first study to examine sex differences for PSVT patients. The primary aim of this study was to investigate if the distribution of potential risk factors differed between men and women with PSVT. Our main finding was that there was a significantly larger proportion of men than women having been involved in strenuous muscular activity preceding debut of PSVT. This means we found a larger incidence of effort-induced PSVT, also known as Paget-Schroetter syndrome (PSS), in men than women. We also found a tendency that women were younger at diagnosis of PSVT compared to men.

Our study, investigating 135 PSVT patients, found an incidence of PSS for both sexes of approximately 58%, which is just between the 50% reported by Aziz et al. and the 60-80% reported in an review by Illig et al. (5,11). Martinelli et al. reported PSS amongst only 25% of their 115 investigated patients (25), while the incidence of PSS in our study was approximately 55% for men and only 32% for women. One study investigating 51 PSVT patients, found 20 effort-induced cases (39%) (26). Among these 20 patients, 15 where men (75%) and 5 where women (25%) (26). There are no certain explanations to the sex-specific proportion of PSS in our study, though a possible interpretation could be that the predominance amongst men may devolve upon a higher participation rate in sports and gym activities amongst men than women. This reasoning is supported by the figures given for attendance for men in “regular work/sports/activity affecting arm”, which roughly corresponds to the figures for strenuous muscular activity in the study; however, there was quite a large proportion of missing values in this variable.

The median age of the patients was 29 years which is consistent with other studies who found the median age for PSVT patients to be around 30 years (5,25). We found no significance difference according to the median age between men and women in our study. However, since the age was a non-normal distributed variable, it was divided by tertiles and we did find a tendency that women were younger than men at PSVT diagnosis in our material. To our knowledge a younger debut age for women compared to men has not been reported by other studies in the field, although Héron et al. reported a younger median age for effort-related UEDVT compared to idiopathic UEDVT (26). However, in our study, age was not analyzed based on effort-induced vs idiopathic PSVT. The tendency we found towards younger PSVT debut for women, could possibly be explained by the fact that young women could have one additional potential risk factor that men are lacking: oral contraceptives (OC). However, it was not investigated exactly how many of the women in the youngest age group took oral contraceptives in our study. Neither has the role of oral contraceptives as a risk factor for PSVT been fully understood. As stated previously Martinelli et al. suggested that OC should not be considered a risk factor unless combined with thrombophilia (25). The prevalence of OC in our study population was about 30%, though it is difficult to draw any conclusions from this since almost 60% of data for this variable was missing.

Among the other investigated variables, we found no significant differences between the sexes. It was more common that the right arm was affected by thrombosis than the left in our study. This finding corresponds with previous studies, likely because of the incidence of right-hand dominance (5). In our study population, there was approximately 18% men and 14% women smoking. When compared to figures from Public Health Agency of Sweden (Folkhälsomyndigheten) on smoking in Stockholm county for men (8-9.9%) and women (9.2-11%), it is obvious that the smoking incidence in our study was higher than in the average population in Stockholm (27). Also, in our study there were more smoking men than women, as opposed to the incidence figures for Stockholm county. However, the difference for smoking between men and women was not significant and it is not possible to draw any conclusions from this since the study design does not include any controls. The frequency of known clotting disorders found in our study population was 3% which is consistent with other studies (17,28). Also, the figure roughly corresponds to the mean prevalence of clotting disorders in European populations (5). The mean BMI was 23.7 in our study population, which is similar to previous studies and indicates that obesity does not constitute a risk factor for PSVT (3,12).

The secondary aim was to examine whether there were any differences regarding time from symptom to diagnosis and treatment between men and women. To our knowledge, there are no such studies in thrombosis patients. A systematic review, examining the subject amongst myocardial infarction patients suggested that patient delay was significantly longer for women than men (29). The majority of studies reviewed found women more likely to delay seeking medical care than men even after adjustment for potential confounders such as age and comorbidity (29). As opposed to that study, we found a strong tendency for longer patient delay in men compared to women in both the acute group (patients treated within 14 days) and in the group consisting all PSVT patients. After adjustment for the variable strenuous muscular activity by a multivariate ANOVA model, this difference remained significant. Our study showed that male patients had longer patient delay than women regardless of preceding strenuous muscular activity. However, our study gives no explanation to this circumstance. One could speculate in socially constructed gender differences contributing to the delayed medical search patterns in men.

Patient delay was shorter among both men and women with preceding strenuous muscular activity, than among patients without. The overall shorter delay amongst patients who had been engaged in strenuous muscular activity before the PSVT debut, could possibly be explained by that symptoms where triggered by an actual physical event which makes the symptoms easier to acknowledge. One can argue that people in general, both men and women, are more likely to seek medical help if debut of symptoms are strongly connected to a physical event, and less prone to seek medical help if the debut of the symptoms is vaguer and arises independently.

A study investigating delay patterns in tuberculosis patients in men and women in Vietnam found both significantly longer patient delay and doctors delay for women (30). Another study investigating tuberculosis in South India, found longer patient delay for men, but longer doctors delay for women (31). Our study showed no significant difference in doctors delay between the sexes. When doctors delay was analyzed with the ANOVA model, an interaction for sex and strenuous muscular activity was found. Doctors delay was longer for men with strenuous muscular activity than men without. For women, the relationship was reversed: women with strenuous muscular activity had shorter doctors delay than women without. One could argue that strenuous muscular activity preceding PSVT debut could be a prolonging factor according to doctors delay, since there is a risk for misdiagnosing the thrombosis as a muscular affliction if symptoms had started after muscular activity. This reasoning would not explain the reversed ratio amongst women though. There are studies showing longer doctors delay for women than for men (30–32). Considering these studies, one possible interpretation could be that women who present a specific triggering factor for their symptoms such as strenuous muscular activity, are more likely to have a shorter way to diagnosis than a woman who presents symptoms without such a factor. However, our study gives no answers on this subject.

### Strengths and limitations

To our knowledge this study is unique since it is the first study investigating potential sex differences in PSVT patients. A major strength of this study was the relatively large sample size and the wide range of collected data on each patient. Also, our cohort ranged through a large year span, from 2007 to 2017. Since this was a retrospective cohort study which included all PSVT patients within a well-defined area, there was no selection bias.

One limitation was the study design; a retrospective study has lower validity than a prospective. Another limitation was the lack of control patients. Without controls it is impossible to fully evaluate the independent risk factors for having PSVT. The aim of this study was instead to study the risk factors, comparing men with women. Furthermore, data were collected manually, which is always a potential source of error, especially when data from medical journals needs to be interpreted to fit defined variables. However, the variables had been defined in advance to facilitate data collection and avert arbitrary interpretation. Since data were population based the results are possible to extrapolate. As with most registry studies there will be missing data, we identified a proportion of missing data, and to be noted was that the largest proportion of missing data was identified amongst women. This was a potential source for confounding bias, although missing data were excluded from the analyzes itself to minimize contamination.

### Clinical significance

There is still some uncertainty surrounding the PSVT diagnosis. What potential risk factors that triggers the disease and who is at risk of getting it remains partly unclear. Our study confirms an anticipated young median debut age. Furthermore, it showed that men had a greater prevalence of effort-induced PSVT than women, and an incidence of almost 60% of PSS in the PSVT population all together. Moreover, the study implied that men had longer patient delay than women. The clinical application for these findings are of great importance. It is crucial for clinicians to recognize and diagnose this rare condition and to rapidly treat it accordingly. Without the knowledge that muscular effort is a common reason for UEDVT, clinicians might misinterpret patient’s symptoms as muscular affliction. The choice of treatment strategy is entirely based on time from symptom debut and longer patient delay might limit the possibility to treat the patient with CDT, since it should be performed within 14 days from onset of symptoms.

### Future studies

More research is needed in the field of PSVT to provide clinicians with information on epidemiology, predisposing factors and optimal treatment of the disease. A study similar to ours but with a matched control group of healthy subjects is needed to give the relative prevalence of risk factors for PSVT in men and women of different ages.

A standardized protocol for patients to fill in at their first health care visit in order to get more information on circumstances of debut, would improve data validity. Strict definitions of strenuous muscular activity i.e. effort-induced PSVT would minimize different interpretation of important events preceding thrombosis. This type of protocol would form a firm basis for more conformational data which is desirable for future studies. A question raised during the work with this study was the potentially larger proportion of missing data amongst women. To investigate whether it could be a systematic tendency that clinicians are more meticulous when taking up patients’ medical history among men than women, a registry study looking in to this matter through patients’ medical journals would be of interest.

## Conclusions

This study found that strenuous muscular activity preceding debut of PSVT was more common among men than women. A majority of the investigated men had effort-induced PSVT, also known as Paget Schroetter syndrome, compared to less than a third of the women. This may be explained by the finding of higher incidence of regular muscular activity involving the arm among men. The study also showed a tendency that women were younger than men at debut of PSVT. Furthermore, a tendency that men had longer patient delay than women was unveiled.

## Data Availability

Individual participant data underlying the results reported in this Article will be made available after de-identification, alongside a data dictionary, study protocol, and informed consent form. The data will be available at Article publication and for 10 years subsequently. Data will be shared for individual participant data meta-analysis with other members of the research community who have an affiliation to a recognised medical university. Data will only be shared with investigator support and after approval of a proposal, and with a signed data access agreement. Additional restrictions apply according to Swedish law.

## Abbreviations

CDT: Catheter-directed thrombolysis
CI: Confidence interval
CHEST: American College of Chest Physicians
CT: Computed tomography
CVC: Central venous catheter
DUS: Duplex ultrasound
DVT: Deep venous thrombosis
IQR: Inter quartile range
MRI: Magnetic resonance imaging
OC: Oral contraceptives
PE: Pulmonary embolism
PSS: Paget Schroetter syndrome
PSVT: Primary subclavian vein thrombosis
PTS: Post-thrombotic syndrome
TOS: Thoracic outlet syndrome
SWEDVASC: Swedish quality registry for vascular surgery
UEDVT: Upper extremity deep venous thrombosis
VKA: Vitamin K antagonist
VTE: Venous thrombus embolism
V-TOS: Venous thoracic outlet syndrome

## Acknowledgements and contributions

I would like to give my warmest thanks to my supervisor Jonas Malmstedt, PhD, MD, for his continuous support and guidance from the beginning to the end of this project. I would also like to thank my future colleague Sara Blomstrand, for great companionship when collecting data together. Finally, I would like to thank my coordinator Per Tornvall and the students in my coordinator group for valuable thoughts and input throughout the writing process of this report.

